# Assessing Ankle Range of Motion with Wearable Technology: A Comparative Accuracy and Reliability Study

**DOI:** 10.1101/2025.05.02.25326892

**Authors:** Miral Atout, Hadil Mahmoud, Mariam Mahmoud, Abdullah Jabri, Shahryar Warsif, Dhruvil Patel, Farhan Raza, Taylor Chomiak, Bin Hu

## Abstract

**Background:** Accurate measurement of ankle range of motion (ROM) is essential for diagnosing and treating musculoskeletal conditions, optimizing athletic performance, and managing neurological disorders such as Parkinson’s disease. This study evaluates the novel Ambulosono device, a sensor-based tool, against the traditional goniometer for assessing Ankle ROM in healthy participants.

**Methods:** A comparative cross-sectional study was conducted on 54 healthy participants aged 15 to 24 years. Ankle ROM was measured using the goniometer, placed on the lateral malleolus, and the Ambulosono device, secured on the dorsum of the foot. Participants performed maximal dorsiflexion and plantarflexion with knees extended, and five measurements were taken per device on both feet in randomized order. Statistical analyses included descriptive statistics, Bland-Altman plots, and Intraclass Correlation Coefficients (ICCs) to assess agreement and reliability.

**Results:** Mean goniometer ROM was 58.44° (SD=5.54) versus Ambulosono’s 56.80° (SD=3.88). No significant differences emerged between devices, foot sides, or gender. Bland-Altman analysis indicated agreement without P proportional bias. Reliability was excellent (Cronbach’s alpha=0.983, ICC=0.983).

**Conclusion:** The Ambulosono device is a robust tool that offers a reliable alternative to traditional goniometry, with advantages such as real-time feedback and reduced inter-rater variability. Its potential applications extend beyond clinical and athletic settings to include neurological rehabilitation and remote patient monitoring. Further research is warranted to validate its efficacy across diverse populations and real-world scenarios.

## Introduction

Ankle injuries are among the most common musculoskeletal complaints, frequently resulting in long-term disability and significant time lost from work or sports. In professional athletes, over 25% of musculoskeletal injuries involve the foot and ankle, with ankle inversion injuries accounting for approximately 50% of all sports-related injuries [1]; in the United Kingdom, ankle sprains represent 3% to 5% of all emergency department visits, equating to roughly 5,600 cases daily. Despite their high prevalence, ankle injuries are often perceived as benign and expected to resolve quickly with minimal intervention. However, this perception overlooks the potential for chronic issues, such as reduced range of motion (ROM) and increased susceptibility to future injuries.

Ankle ROM is a critical movement for various activities, such as walking, running, and jumping [2]. Accurate measurement of this range of motion (ROM) is essential for diagnosing and treating a variety of conditions, as well as for assessing athletic performance [3].

The goniometer—a hinged device with two arms that can be aligned with anatomical landmarks—has been traditionally employed to measure Ankle ROM due to its simplicity, cost-effectiveness, and ease of use [4]. Despite its widespread application, the accuracy of goniometric measurements is subject to several influencing factors, including the examiner’s experience, the patient’s cooperation, and the specific technique employed [4]. The manual handling of the patient’s limb inherent in goniometry can introduce further variability. These limitations can pose significant challenges, particularly when precise measurements are required for clinical decision-making or research purposes [5].

One of the main limitations of traditional measurement tools like the goniometer is their inability to measure dynamic data [6]. They are designed to measure static angles—i.e., the position of a joint at a specific point in time—and cannot capture the continuous changes in joint angles that occur during movement [6]. This limitation can be a significant issue in fields like physiotherapy and sports science, where understanding the dynamics of joint movement is crucial [5].

To address these limitations, various devices have been developed to improve the accuracy and reliability of Ankle ROM ROM measurements. The Ambulosono device is a sensor-based measurement tool that offers a potential solution to these challenges [7,8]. This wearable sensor-based system is designed for biofeedback training and represents a novel approach to measuring Ankle ROM [7,8]. The device uses electromyographic signals and a biomechanical model of the lower leg to estimate joint stiffness during dorsiflexion and plantar flexion [7,8].

The Ambulosono device can also mitigate psychological factors by allowing patients to perform the tests in their environment, at their own pace, and without an observer [9]. This can significantly reduce the potential for anxiety or discomfort that could affect the patient’s performance [9]. Furthermore, by eliminating the need for an observer, the Ambulosono device also removes the potential for inter-rater and intra-rater variability, which can further improve the accuracy and reliability of the measurements [8,9].

Other devices have also been developed to measure ankle ROM. The WIMU, for instance, is a wearable inertial measurement unit that has been used to measure Ankle ROM [10]. It has shown high reliability and validity when compared with a standard goniometer and a 2D video-based motion analysis software [10]. However, the WIMU requires placement on the leg, unlike the Ambulosono device that can be attached adjacent to the focused joint, which could potentially affect the accuracy of measurements [10]. Furthermore, it does not provide real-time feedback, which is a significant advantage of the Ambulosono device [10].

Several devices are conventionally employed to measure ankle range of motion (ROM), each with its own set of limitations. Traditional inclinometers, often used to assess weight-bearing ankle ROM, provide a measure of the tibia’s inclination relative to the ground [11]. In recent years, smartphone inclinometer applications have emerged as a convenient alternative, offering digital displays and ease of use [12]. The D-Flex, a newer device, offers a standardized method of ankle fixation and an automatic measurement system, thereby reducing potential human error [11].

The development and validation of accurate, reliable, and user-friendly devices for measuring Ankle ROM are crucial for both clinical and research settings. The Ambulosono device, with its ability to provide real-time feedback and capture dynamic movements, could significantly contribute to this field [7]. However, further research is needed to assess its accuracy and reliability compared to other measurement methods or devices.

## Methods

### Study design

This study utilized a comparative cross-sectional design to assess the accuracy of a unique instrument, the Ambulosono device, in measuring Ankle ROM compared with a typical goniometer. Prior to data collection, raters received comprehensive training on the proper use of the goniometer. This training had two aspects: first, raters viewed a detailed video created by a specialist in the field on accurately measuring Ankle ROM with a goniometer; second, raters practiced with each other to improve their skills.

### Participants

A total of 54 university students (16–24 years of age) participated in various subprojects of this study. This sample size is comparable to that used in previous studies [13,11]. All participants provided written consent and formal registration prior to data collection. They were free from injury, had no history of hospitalization or chronic diseases affecting exercise capacity, were not involved in competitive sports, and were lifelong non-smokers. Ethics approval was obtained from the University of Calgary Research Ethics Board as part of the Ambulosono registered trial (ISRCTN06023392).

### Instruments

The Ambulosono device is a single sensor-based measurement tool [7–9], comprising 3-axis MEMS-based gyroscopes and accelerometers with validated accuracy. The Ambulosono app employs fusion algorithms for automatic gravity calibration and real-time angular velocity output (pitch, roll, yaw), providing accurate ROM measures. The App samples sensor output at 50 Hz based on the angular velocity produced by joint rotations and stored on the smartphone before being uploaded to an encrypted data server. The data collected via Ambulosono device was compared to that obtained by the goniometer [4], which were manually collected and recorded.

### Procedure

Participants were instructed to lie supine on a level surface with their feet in a resting position. They were then instructed to dorsiflex as close to 90 degrees as possible to obtain the starting point (the “zero position”), which was used as a reference line to which participants reset after each ankle flexion. Once this was accomplished, participants were asked to perform maximum Ankle ROM, and the raters manually recorded the values revealed by the goniometer—placed on the lateral malleolus with the stationary arm aligned along the lateral side of the leg and the moving arm parallel to the foot [Figure 1]. Next, participants were asked to perform maximum plantarflexion, and values were re-recorded. This was repeated five times on each foot.

**Figure 1.**
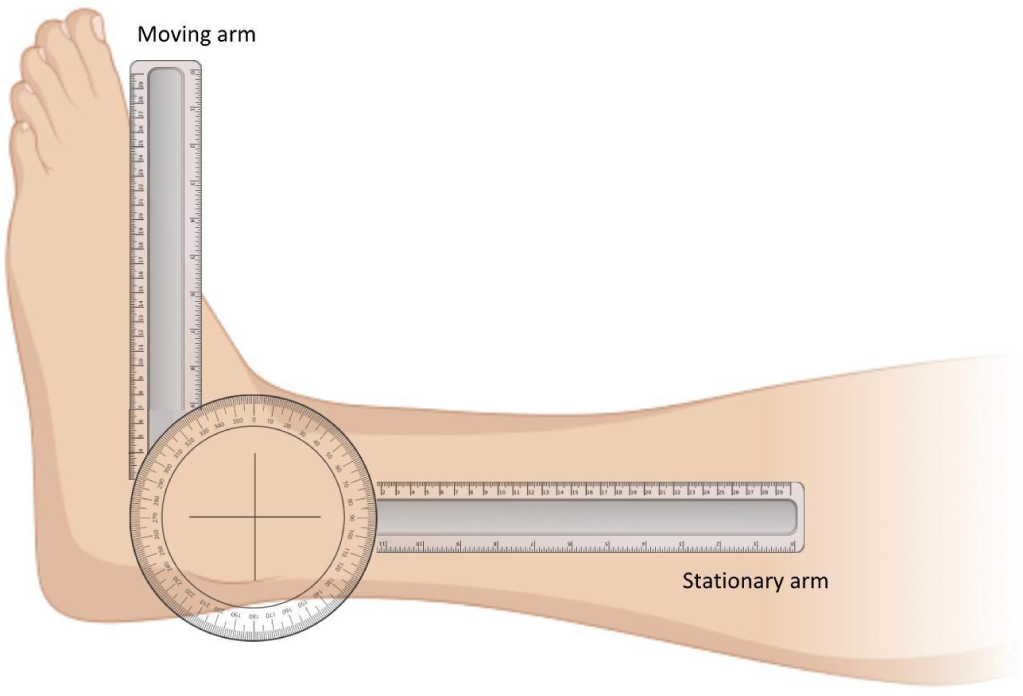
Goniometer placement for measuring Ankle ROM. The stationary arm is positioned along the lateral midline of the leg, and the moving arm is aligned parallel to the foot. The axis of the goniometer is centered over the lateral malleolus to capture the ankle’s range of motion.

The same procedure was then repeated with the Ambulosono device, which was fixated within a glove on a brace that was wrapped around the dorsum of the foot [Figure 2].

**Figure 2.**
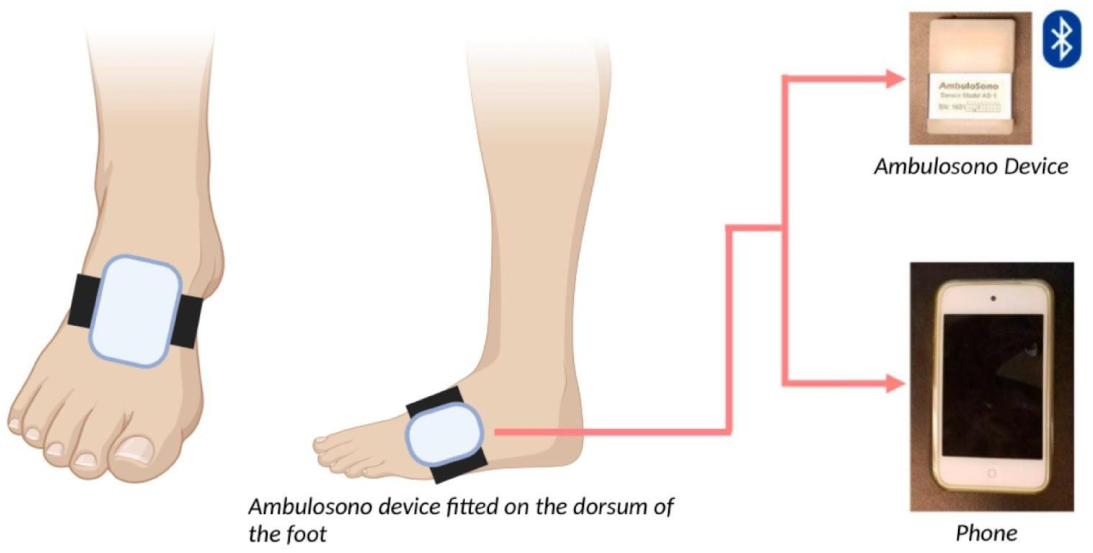
Ambulosono device positioning and data flow. The wearable sensor is fitted on the dorsum of the foot and transmits motion data via Bluetooth to a smartphone for real-time monitoring and recording of Ankle ROM.

While participants performed dorsiflexion and plantarflexion, the sensor automatically calculated and recorded the data obtained with each movement. This was done continuously for 10 seconds.

### Statistical Analysis

Agreement between the Ambulosono device and the goniometer data was analyzed using python codes on OpenAI advanced data platform and with SPSS version 29. Cross validations were obtained between the two softwares. Statistical tests included descriptive analysis, the Kolmogorov-Smirnov Test, the Intraclass Correlation Coefficient (ICC) and Bland-Altman analysis [14,15].

## Results

### Goniometer Data

Figure 3 shows the distribution of ankle ROM derived from the sum of its components (dorsiflexion and plantarflexion) measured using a goniometer. The data were collected from thirty-seven participants, who had both goniometer and Ambulosono device measurements taken for the left and right foot.

**Figure 3:**
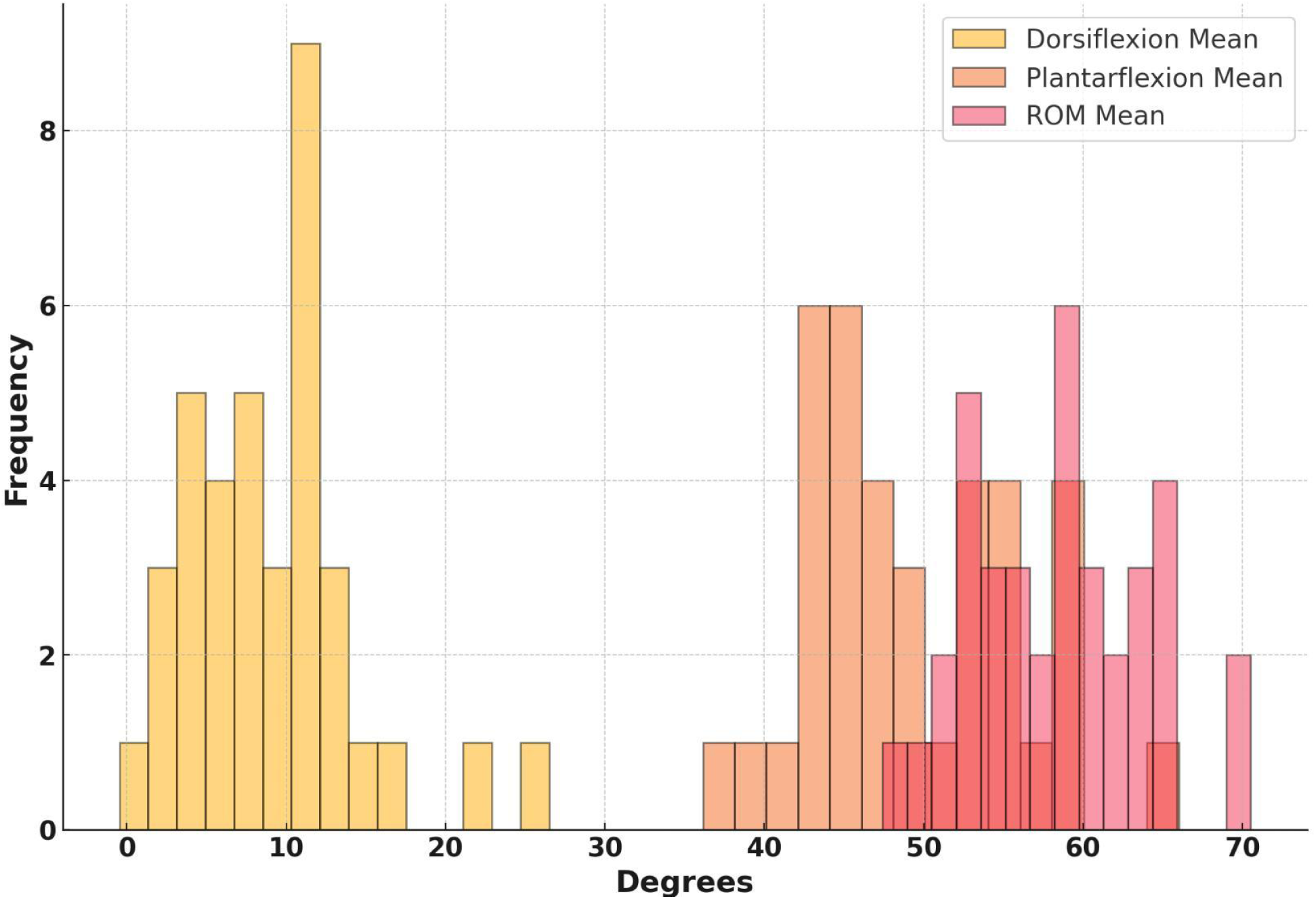
Frequency distribution of dorsiflexion, plantarflexion, and range of motion (ROM) means measured using a goniometer. The histogram highlights the distinct ranges for dorsiflexion (yellow), plantarflexion (orange), and the combined ROM (red), illustrating the relative contributions of each movement to the total ROM.

### Ambulosono device

The Ambulosono device was used to measure the ROM during continuous ankle rotations. Unlike a goniometer, the Ambulosono device does not have a defined zero point and cannot differentiate between plantarflexion and dorsiflexion. Instead, it provides a single aggregated ROM measurement, making it useful for dynamic assessments where distinguishing between specific motions is not required.The Shapiro-Wilk normality test shows that the Goniometer data is normally distributed (p = 0.591), but the Ambulosono data is slightly non-normal (p = 0.082). However, the p-values suggest near-normality for both.

### Descriptive statistics

The goniometer data (Table 1) shows that dorsiflexion has a mean of 9.06 degrees (SD = 5.34) and ranges from −0.46 to 26.50 degrees, while plantarflexion has a mean of 49.38 degrees (SD = 6.61) with a range from 36.15 to 66.00 degrees. The ROM measured by the goniometer has a mean of 58.44 degrees (SD = 5.54), ranging from 47.42 to 70.52 degrees. In comparison, the Ambulosono device measures ROM with a slightly lower mean of 56.80 degrees (SD = 3.88), ranging from 46.43 to 64.02 degrees. Statistical tests indicate that both goniometer ROM and Ambulosono ROM data are normally distributed, as determined by the Shapiro-Wilk test (p > 0.05). Furthermore, an independent t-test revealed no statistically significant difference in the value frequency distributions of ROM between the two devices (p = 0.1442). These results highlight comparable measurements between the two devices, with slight differences in precision and variability.

**Table 1:** Descriptive statistics for dorsiflexion, plantarflexion, and ROM measured using a goniometer, alongside ROM measured using the Ambulosono device. The Ambulosono ROM column is highlighted in green to distinguish its aggregated measurement from the goniometer-derived values.

|  | **Dorsiflexion**<br>**(Goniometer)** | **Plantarflexion**<br>**(Goniometer)** | **ROM**<br>**(Goniometer)** | **ROM**<br>**(Ambulosono Device)** |
| --- | --- | --- | --- | --- |
| Mean | 9.06 | 49.38 | 58.44 | 56.8 |
| Std Dev | 5.34 | 6.61 | 5.54 | 3.88 |
| Min | -0.46 | 36.15 | 47.42 | 46.43 |
| 25% | 5.83 | 44.92 | 54.0 | 55.12 |
| 50% | 8.79 | 48.0 | 58.6 | 57.44 |
| 75% | 11.12 | 54.33 | 62.54 | 59.78 |
| Max | 26.5 | 66.0 | 70.52 | 64.02 |

### Comparative analysis

The Bland-Altman plot in Figure 4 was used to evaluate the agreement between the ROM measurements obtained using the goniometer and the Ambulosono device. The plot revealed a small mean bias, indicating a close agreement between the two devices. The limits of agreement, calculated as the mean difference ±1.96 standard deviations, showed that most data points fell within this range, suggesting consistent measurements across participants. The distribution of differences did not exhibit proportional bias, as there was no apparent trend between the mean ROM values and the differences. These findings highlight that while the Ambulosono device differs in methodology by providing aggregated ROM values without differentiating plantarflexion and dorsiflexion, it demonstrates comparable performance to the goniometer in measuring total ROM during continuous ankle motion.

**Figure 4.**
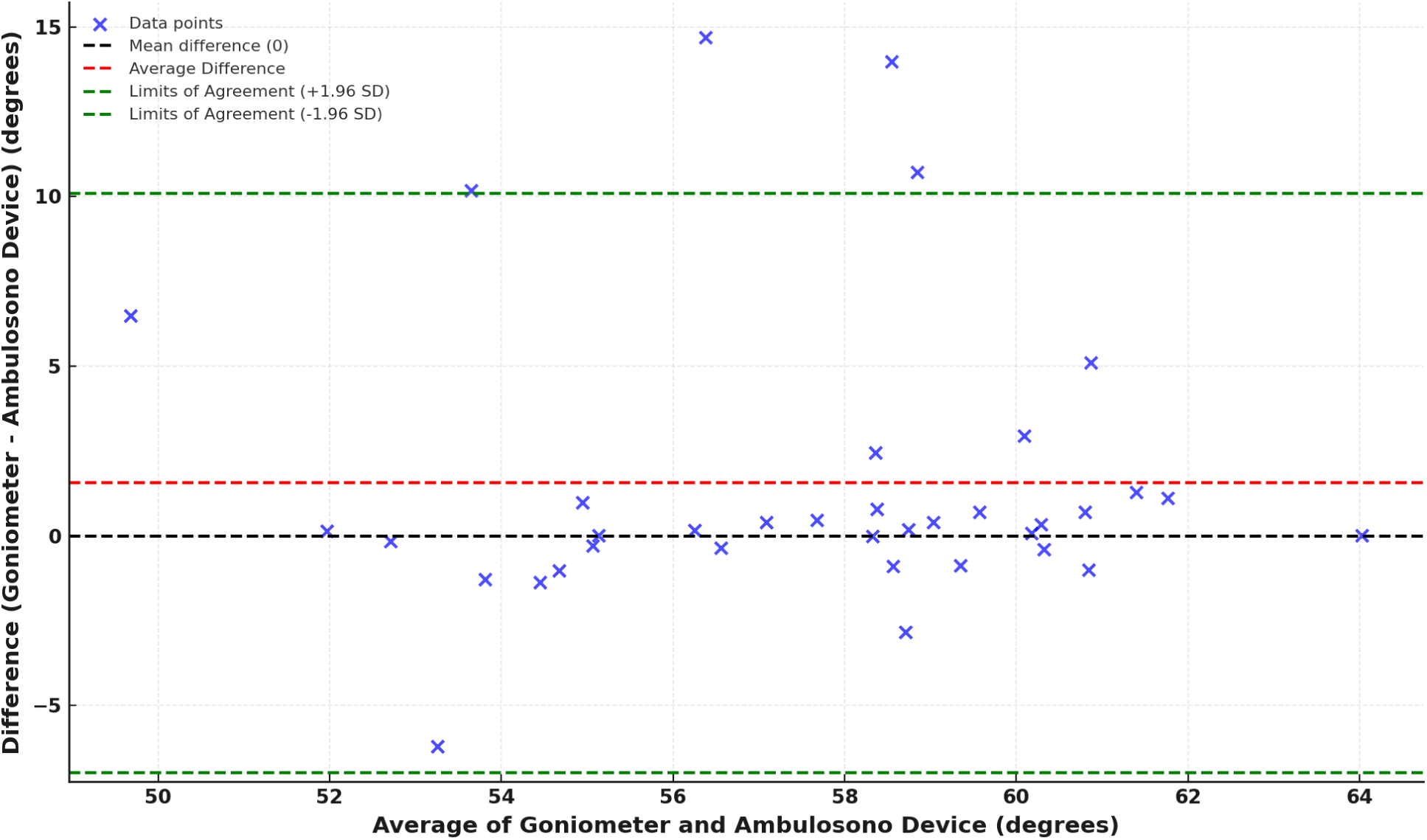
Bland–Altman plot comparing goniometer and Ambulosono measurements of Ankle ROM. The x-axis shows the mean of the two methods, while the y-axis shows their difference (goniometer minus Ambulosono). The central dashed (blue) line represents the mean difference, and the upper and lower dashed (red) lines mark the 95% limits of agreement. Each yellow “×” denotes an individual data point.

**Figure 4:**
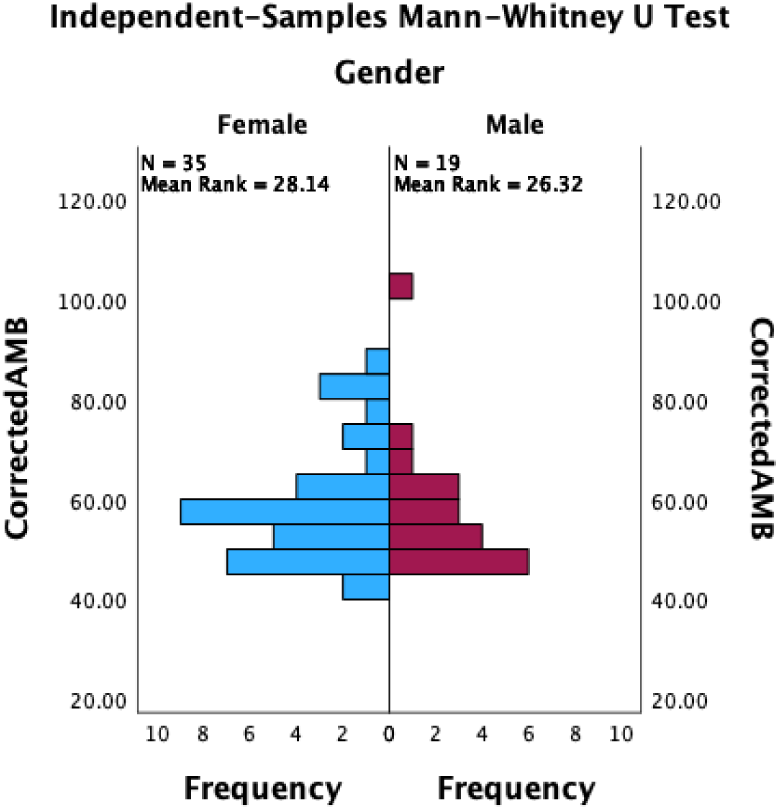
Mirrored histogram of Ambulosono device (correctedAMB) measurements obtained from 54 subjects separated by female (blue bars) and male (magenta bars). N indicates the number of participants in each group, and “Mean Rank” is derived from the Mann–Whitney U analysis. Despite minor differences in distribution, the mean ranks (28.14 for females vs. 26.32 for males) suggest no statistically significant difference between the two groups.

**Figure 5:**
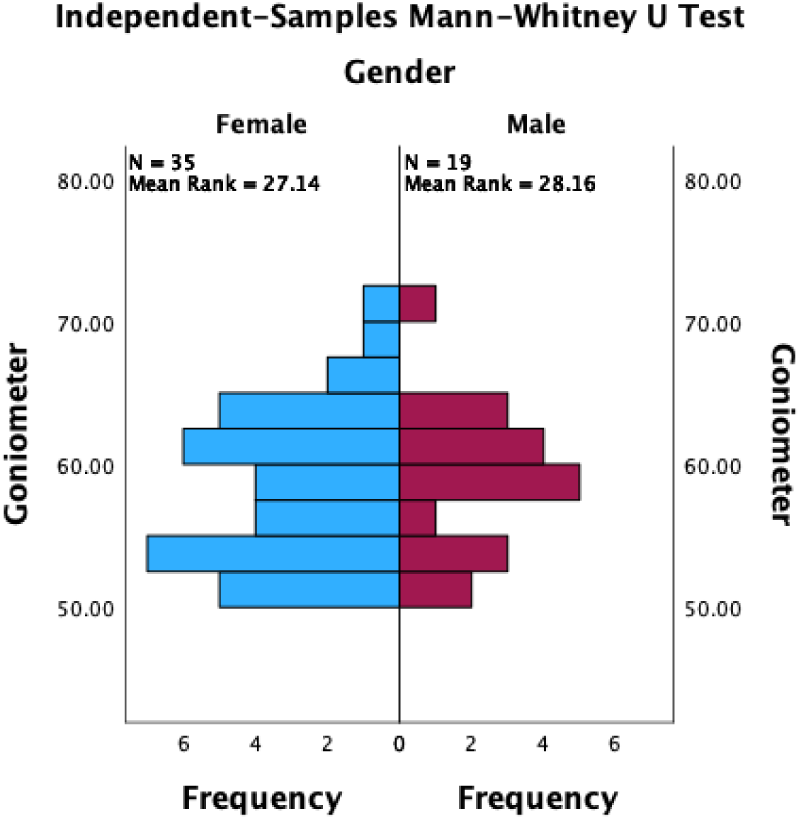
Mirrored histogram of goniometer plantarflexion measurements between female (blue bars) and male (magenta bars) participants. Sample sizes (N=35 for females, N=19 for males) and their respective mean ranks (27.14 vs. 28.16) appear above each distribution. The y-axis denotes plantarflexion values, while frequency is shown along the x-axis. The close similarity in mean ranks suggests no statistically significant difference in goniometer measurements between the two groups.

**Figure 6.**
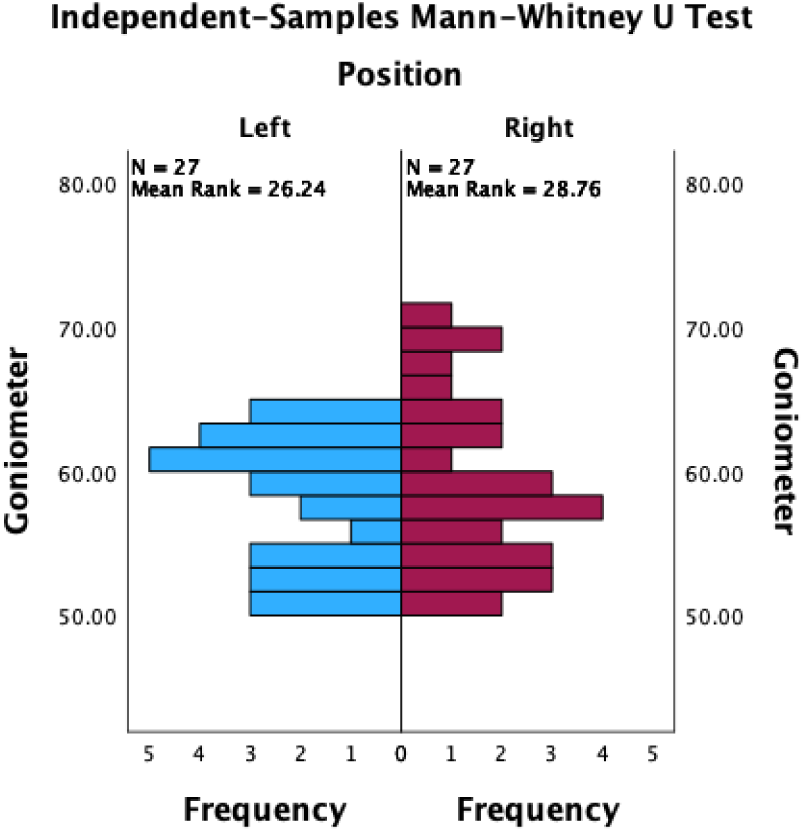
Mirrored histogram of goniometer plantarflexion measurements with an independent-samples Mann–Whitney U test comparing left foot (blue bars) and right foot (magenta bars). Both groups contain N = 27 participants, with mean ranks of 26.24 (left) and 28.76 (right). The y-axis displays goniometer angles, and the x-axis shows frequency. The minimal difference in mean ranks suggests no statistically significant difference between left- and right-foot measurements.

**Figure 7.**
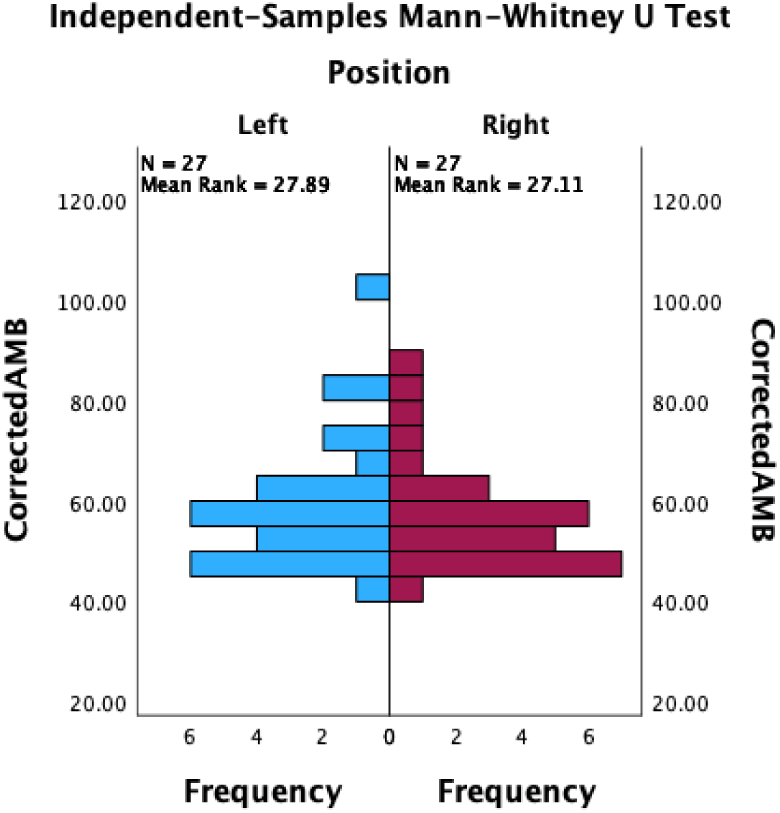
Mirrored histogram of Ambulosono measurements with an independent-samples Mann–Whitney U test comparing the left foot (blue bars) and the right foot (magenta bars). Each group contains N = 27 participants, with mean ranks of 27.89 (left) and 27.11 (right). The y-axis represents Ambulosono values, while frequency is displayed on the x-axis. The similar mean ranks indicate no statistically significant difference between left- and right-foot plantarflexion.

Furthermore, neither goniometer nor Ambulosono measurements differ significantly by gender or by foot (position). All p-values exceed 0.05, so the null hypotheses are retained in every case. In other words, the Mann–Whitney U tests show no statistically significant difference in angle measurements (corrected or uncorrected) across gender or between the left and right foot. The corresponding Kolmogorov–Smirnov tests confirm no difference in the distributions of the measurements across these same categories, corroborating findings in other studies [18–20].

Reliability between two raters was assessed using Cronbach’s Alpha and kappa statistics. These reliability statistics show an overall Cronbach’s alpha of 0.983, indicating excellent internal consistency between the two raters. The Intraclass Correlation Coefficient for average measures is likewise 0.983 (95% CI: 0.972–0.990), reflecting excellent agreement. The inter-item correlation matrix (0.969) further underscores the high level of consistency. Collectively, these values demonstrate that measurements from the two raters are highly reliable and consistent with one another, suggesting near-perfect agreement [15].

## Discussion

The ankle joint, a hinged synovial joint formed by the articulation of the talus, tibia, and fibula bones, serves as a critical kinetic link between the lower limb and the ground. This connection is essential for walking and other weight-bearing activities. Two primary movements of the ankle occur at the tibiotalar joint in the sagittal plane: dorsiflexion and plantarflexion. Dorsiflexion involves bending the foot toward the shin, decreasing the angle between them, while plantarflexion refers to pointing the foot downward by lifting the heel. These movements are fundamental to mobility and are often compromised following ankle injuries.

ROM is a critical factor in joint health and injury risk. Limited dorsiflexion ROM, for example, has been linked to an increased risk of knee-valgus displacement and potential ACL injuries [1]. In individuals with Parkinson’s disease, ROM limitations are further exacerbated by rigidity, bradykinesia, and postural instability, severely impairing gait and balance [9].

Our findings reveal that the Ambulosono device exhibits wider variability (SD = 12.20) compared to a traditional goniometer (SD = 5.14). This difference underscores the inherent measurement approaches of each device. The goniometer captures a single, static value, whereas the Ambulosono device continuously measures multiple angles, reflecting dynamic and potentially more “real-world” behavior. Such continuous capture offers valuable insight into subtle variations or outliers that static methods can overlook [6,16].

Bland-Altman plots showed a mean difference near zero between the two methods, but with broader limits of agreement for the Ambulosono device [14]. While wider, this range suggests greater sensitivity in capturing dynamic changes, an attribute advantageous in both clinical and athletic settings. Indeed, any disparities in measurement outcomes can be largely attributable to nature of the measurement methods. When using the goniometer, subjects performed isolated maximal dorsiflexion and plantarflexion movements that were discrete and easier to reproduce, thus minimizing discrepancies caused by such factors as fatigue within each repetition. In contrast, the Ambulosono device was used during continuous movement cycles, capturing both submaximal and maximal angles as well as intermediate rest phases. Consequently, any fatigue or minor inconsistencies over time became embedded in the Ambulosono dataset, creating broader variability and highlighting the real-time fluctuations in ankle range of motion.

Despite the higher variability, the Ambulosono device is highly reliable. By automating data capture and eliminating many user-related errors associated with manual goniometry, it aligns with advancements in wearable tech [21,23], reduces inter-rater variability, and offers real-time feedback. It also allows patients to take measurements independently, potentially increasing access to consistent monitoring [9].

Additionally, no significant differences were detected between genders or limbs, aligning with prior research [18–20]. This supports the device’s broad applicability across diverse populations. In athletic contexts, the Ambulosono device could aid in performance monitoring and injury prevention [1]. In rehabilitation, especially post-injury or in neurological conditions, continuous ROM tracking may better guide individualized therapeutic programs.

Although our study focused on healthy cohorts, substantial advanceme 17] have been made in wearable ankle measurement devices in injured patients. For example, the development of robotic platforms for ankle rehabilitation has been a significant focus, with advancements in customized training programs and real-time feedback systems that enhance patient engagement and recovery efficiency (25, 27). These platforms, although promising, face challenges such as high costs and technical limitations, necessitating the development of more affordable and adaptable solutions (25, 27).

Artificial intelligence (AI) has also the potential in identifying predictors of ankle sprains and improving rehabilitation outcomes (26, 28). AI techniques, particularly those utilizing electromyography and joint angle data, have shown high accuracy in injury detection, although challenges remain in sensor design and AI model integration (26). Parallel ankle rehabilitation robots (PARRs) have been extensively studied, with research focusing on optimizing mechanical designs and control strategies to improve rehabilitation effectiveness (27, 31). However, the need for clinical trials to validate these technologies remains a critical step for their integration into clinical practice (31). The use of wearable technology, such as inertial measurement units (IMUs) and series elastic actuators, has been highlighted for its potential in providing cost-been the and efficient rehabilitation solutions (28, 29). These devices offer the advantage of enabling in-home rehabilitation and providing real-time feedback, which can lead to more consistent therapy outcomes (28, 29). Additionally, controlled ankle motion (CAM) boots have been reviewed for their biomechanical effectiveness in restricting ankle range of motion and redistributing plantar pressure, although further research is needed to fully understand their impact on rehabilitation (30). Overall, these studies underscore the ongoing advancements and challenges in the field of wearable ankle rehabilitation technology, emphasizing the need for continued research and development to enhance clinical outcomes and accessibility (25–34).

## Limitations

First, our sample consisted of healthy participants, which may not reflect the range of motion challenges in clinical populations such as patients with Parkinson’s disease or significant musculoskeletal impairments. Second, the reliance on a single measurement session constrains our ability to assess intra-rater reliability fully. Repeated sessions and more complex tasks—like squatting or jogging—would provide additional insight into the device’s robustness. Lastly, because this wearable technology provides immediate numerical outputs, clear guidelines for interpreting those data are crucial to avoid misapplication in clinical or home-based settings.

## Conclusion

The Ambulosono device offers a reliable alternative to traditional goniometer for measuring ankle ROM. Its capacity for continuous data capture, real-time feedback, and reduced inter-rater variability make it a compelling option in both clinical and athletic environments. Owing to its dynamic measurement capabilities, it holds promise for applications ranging from injury prevention to neurological rehabilitation. Further research with diverse populations and activity protocols is recommended to confirm its broader utility.

## Data Availability

All data produced in the present study are available upon reasonable request to the authors

## Funding Source and Acknowledgement

This study was funded by the Alberta Ministry of Mental Health and Hotchkiss Brain Institute, Cumming School of Medicine, University of Calgary.

